# A Prediction model for Pulmonary Embolism in Patients with Spontaneous Intracerebral Hemorrhage

**DOI:** 10.1101/2024.10.16.24315633

**Authors:** Yuanyun Chen, Xiaoshu Wang, Quanhong Shi, Yanfeng Xie, Yulong Xia, Jianjun Zhong, Li Jiang

## Abstract

**Background and Purpose:** Spontaneous intracerebral hemorrhage (sICH) patients were susceptible to pulmonary embolism (PE), which is frequently neglected due to the absence of predictive tools for early detection. This study aimed to investigate the risk factors of PE in sICH patients and develop a efficient model for the prediction of PE in clinical practice.

**Materials and methods:** We conducted a retrospective study involving 1129 sICH patients. The enrolled patients were divided into training set, internal validation set and external set (n=525: 357: 247). The univariate and multivariate stepwise logistic regression analyses were employed to screen the independent risk factors of the PE in sICH patients. A nomogram model (Model P-P) was constructed based on R language and subsequently validated. For further evaluation, the Model P-P was compared with another similar model (Model A).

**Results:** The analyses revealed that deep vein thrombosis, age, Glasgow coma scale score, fibrinogen degradation product, D-dimer, hemoglobin, hemorrhage volume, plasma osmolality, and surgical method were significant risk factors for PE in sICH patients (p<0.05). The Model P-P was established based on these factors, and its sensitivity, specificity were 84.2%, 94% and 0.910, respectively. Furthermore, the AUC of Model P-P (0.894) was significantly higher than that of Model A (0.785) (p<0.05).

**Conclusion:** The Model P-P developed in this study exhibited reliable predictive efficiency, making it an applicable tool for the early evaluation and prediction of PE after sICH surgery. This model is potential to assist personalized treatment decisions in clinical practice, especially in the neurosurgical intensive care unit.

## Introduction

Spontaneous intracerebral hemorrhage (sICH) is a common emergency in neurosurgery. The prevalence of sICH is approximately 60-80 instances per 100,000 individuals annually, including around 30% of all stroke cases.^1^ It is distinguished by its high fatality rate and propensity for causing disability.^2, 3^ Pulmonary embolism (PE) is globally the third most frequent acute cardiovascular syndrome behind myocardial infarction and stroke.^4^ Patients with sICH have an increased vulnerability to PE due to decreased physical activity resulting from prolonged bed rest, as well as an elevated tendency for blood clot formation (hypercoagulability).^5^ Previous study indicated that as much as 75% of patients with residual hemiplegia after sICH developed deep vein thrombosis (DVT) if they were not received appropriate prophylactic treatment, and PE-related mortality occurs in nearly 5% of sICH patients.^6^ However, PE is often neglected due to its subtle clinical symptoms and the lack of early predictive tools.^7^ Hence, it is highly important to promptly identify the risk factors for PE after sICH surgery. In this study, we launched a retrospective analysis based on our clinical data and developed a new nomogram model with the purpose of assisting early prediction of PE in clinical practice.

## Methods

### Patient population

A continuous cohort consisting of 1,215 sICH patients was established, including those admitted between January 2017 and May 2024. The inclusion criteria were as follows: ① cranial and pulmonary CT scans performed within 24 hours of onset; ② all enrolled patients accorded with the diagnostic criteria for sICH, and underwent surgical treatment within 3 days of onset;^8^ ③ age over 18 years old. Exclusion criteria included: ① heart failure; ② presence of PE, varicose veins, phlebitis, chronic venous insufficiency, and other venous related diseases; ③ coagulation dysfunction and abnormal bleeding in other parts of the body; ④ hematological system diseases; ⑤ previous history of tumor; ⑥ incomplete clinical data.

A total of 1,129 individuals were finally included in the study based on the inclusion and exclusion criteria. Enrolled patients were randomly separated into training set (n= 525), internal validation set (n= 357), and external validation set (n= 247). The study received approval from the Ethics Committee of the First Affiliated Hospital of Chongqing Medical University (K2023-581). The use of non-patient privacy medical records in the study was fully explained to the patient or legal representative during admission, and informed consent and full written authorization were obtained. Computed tomographic pulmonary angiography (CTPA) was performed in all patients to detect the presence of PE by evaluating the distribution and structure of the pulmonary vasculature. A positive CTPA report, as interpreted by the same team of senior radiologists, was deemed the most conclusive and dependable evidence for confirming the diagnosis of PE.^9^

### The relevant risk factors of PE after sICH surgery

According to existing literatures and current related guidelines,^10,11^ the possible risk factors that may impact PE after sICH surgery were screened out for evaluation. These factors included the patient’s gender, age, body mass index (BMI), smoking history, presence of diabetes mellitus, presence of hypertension, Glasgow Coma Scale (GCS) score, presence of hemiplegia, surgical method employed, postoperative conditions (such as ventilator dependence, mechanical prophylaxis, central venous cannulation, tracheotomy, and DVT), presence of systemic infection, blood transfusion, and various laboratory test results (hemoglobin levels, platelet count, mean platelet volume, prothrombin time (PT), activated partial thromboplastin time (APTT), fibrin degradation products (FDP), D-dimer(DD), and plasma osmolality).

### The development process of the PE prediction model for sICH patient

In the training set, the ROC curves were generated using R language (version 4.4.0) to determine the optimal cutoff values,^12, 13^ which allowed for the conversion of quantitative data into categorical data. Initially, univariate logistic regression analysis was conducted to identify risk factors for PE after sICH surgery. These identified risk factors were then subjected to multivariate logistic regression analysis. Subsequently, the independent risk factors that demonstrated statistical significance in the multivariate analysis were utilized to construct a predictive model for PE after sICH surgery. The nomogram model, namely PE prediction model for sICH patient (Model P-P), was constructed using R language. In R language, the dataset was initially prepared through appropriate data reading and cleaning procedures to ensure data quality and integrity. In light of the study’s objectives and characteristics, Binary Logistic Regression Model was deemed the optimal selection for the construction of a predictive model. Following model development, the rms package was employed to develop the nomogram. The datadist function was first used to prepare the data distribution information, and the nomogram function was utilized to create the nomogram model. Finally, the nomogram was plotted using regplot function. Model performance was assessed by sensitivity, specificity, receiver operating characteristic (ROC) curve, and area under the curve (AUC). Additionally, the calibration curves were used to assess the model’s predictive accuracy, and decision curve analysis (DCA) was employed to evaluate its clinical utility.^14-17^ A value of P< 0.05 was considered statistically significant. Figure 1 displayed the flow chart of the investigation.

**Figure 1.**
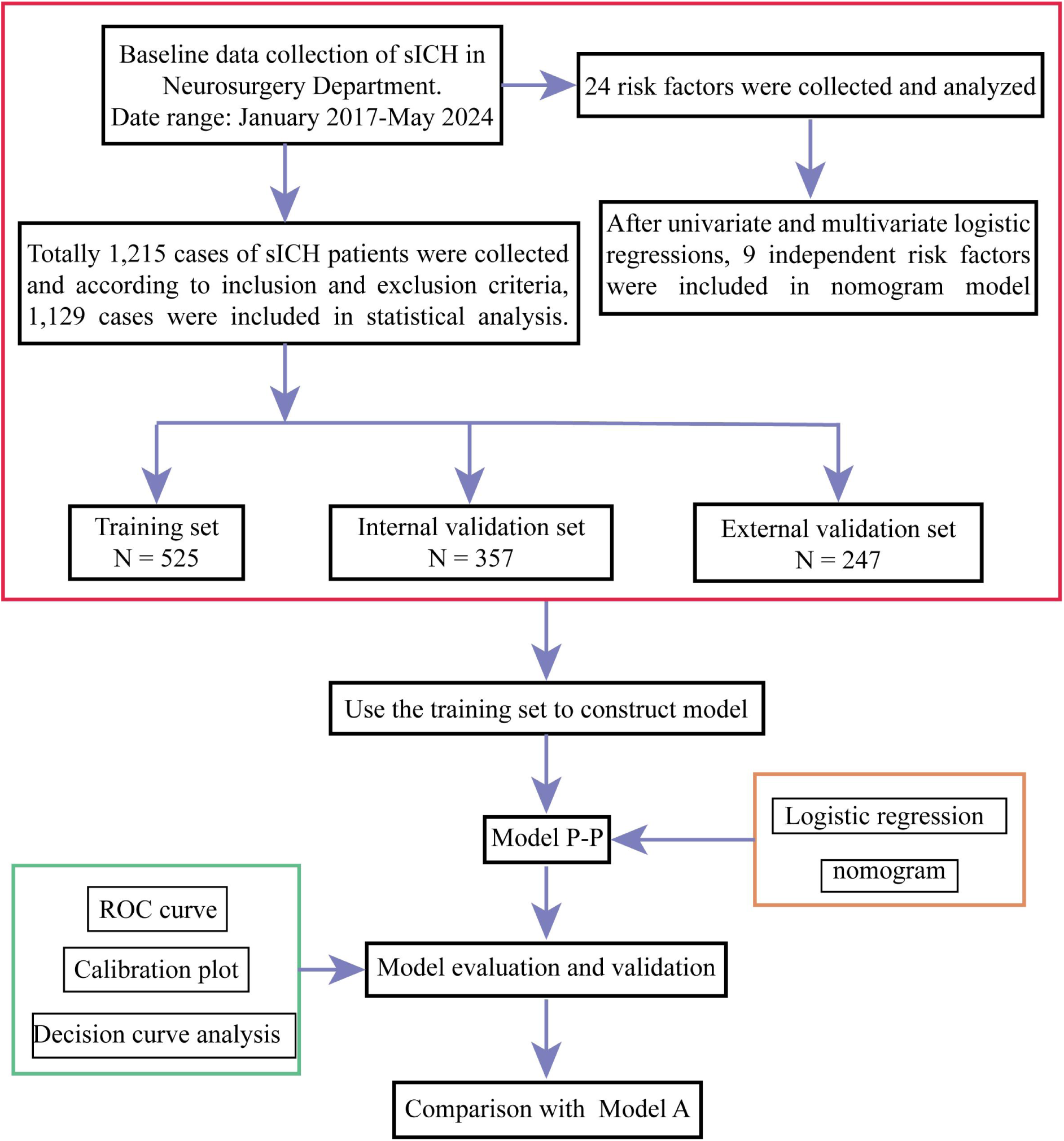
The flow chart of the study. ROC, Receiver Operating Characteristic.

### Comparison with the similar model

In previous study, researchers established a risk prediction model (Model A) for PE after sICH surgery.^18^ To further evaluate the clinical efficacy of our approach, a comparison was conducted between the Model P-P and Model A. The comparative analysis of the two models was conducted using additional 50 sICH patients that were collected at our institution during the period from January to June 2024, in accordance with the inclusion and exclusion criteria. The ROC curve was then plotted based on the total score to compare the discriminatory abilities of the two models.

### Statistical analysis

The statistical analysis was conducted using SPSS 27.0. Variables with a normal distribution were represented as the mean ± standard deviation (x ± s), while non-normally distributed data were represented by the median and percentile [M (Q1, Q3)]. Independent samples t-tests were used to compare sets of variables with normal distribution, and the rank sum tests were employed for non-normal distribution data. Comparisons between groups of count variables were conducted using the χ2 test. ROC curves were generated with the ggplot2 and pROC packages to evaluate the accuracy of the nomogram. Calibration of the prediction model was assessed using calibration curves produced by the ggplot2 package, and clinical net benefit was evaluated through DCA using the ggscidca package.

## Results

### Study design and population

The overall incidence rate of PE after sICH surgery was 11.34%. Specifically, the training set had 57 cases of PE, the internal validation set had 45 cases of PE, and the external validation set had 26 cases of PE. The training set was divided into two groups: PE group and no-PE group. Table 1 and Figure 2 displayed the statistics of the PE group and the no-PE group within the training set. The ideal cutoff value and the univariate analysis results of factors influencing PE after sICH surgery were presented in Table 1. We included variables with P <0.05 in the multivariate logistic regression to confirm independent risk factors. These factors included DVT, age, GCS score, FDP, DD, hemoglobin, hemorrhage volume, plasma osmolality, and surgical method, as presented in Table 2.

**Figure 2.**
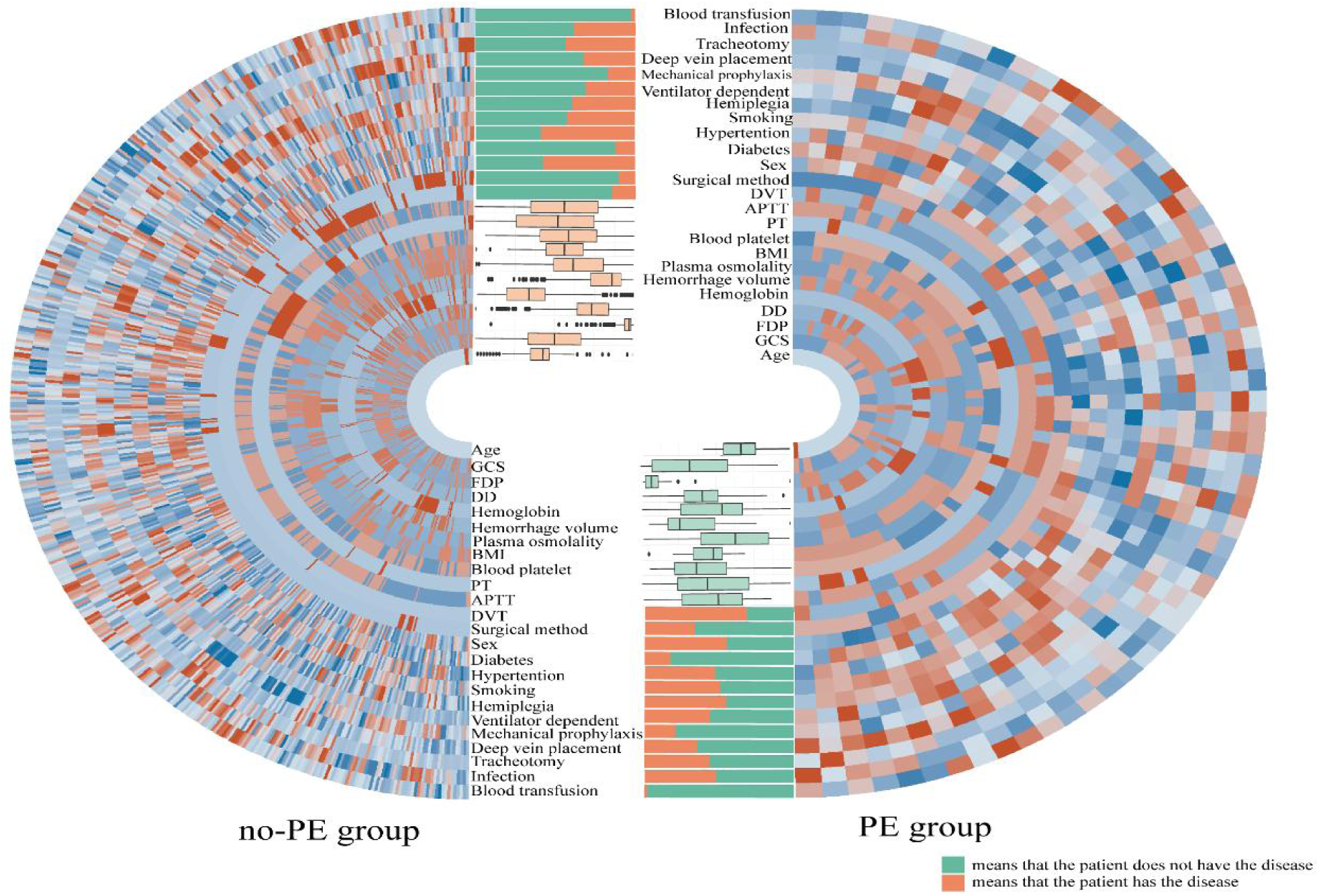
The data distribution chart of PE-related risk factors. The heatmap, boxplot, and bar chart for the PE group and no-PE group. The heatmap showed the distribution of the data values. A closer proximity to red indicated larger values, while a closer proximity to blue indicated smaller values. The box plot illustrated the median and percentile of the training set. The bar chart illustrated the proportion of the categorical data within the training set. The data distribution chart was helpful for screening independent risk factors for PE after sICH surgery. PE, pulmonary embolism; sICH, spontaneous intracerebral hemorrhage; BMI, Body mass index; GCS, Glasgow Coma Scale; FDP, Fibrin degradation products; DD, D-dimer; PT, Prothrombin time; APTT, Activated partial thromboplastin time; DVT, Deep vein thrombosis.

**Table 1.**
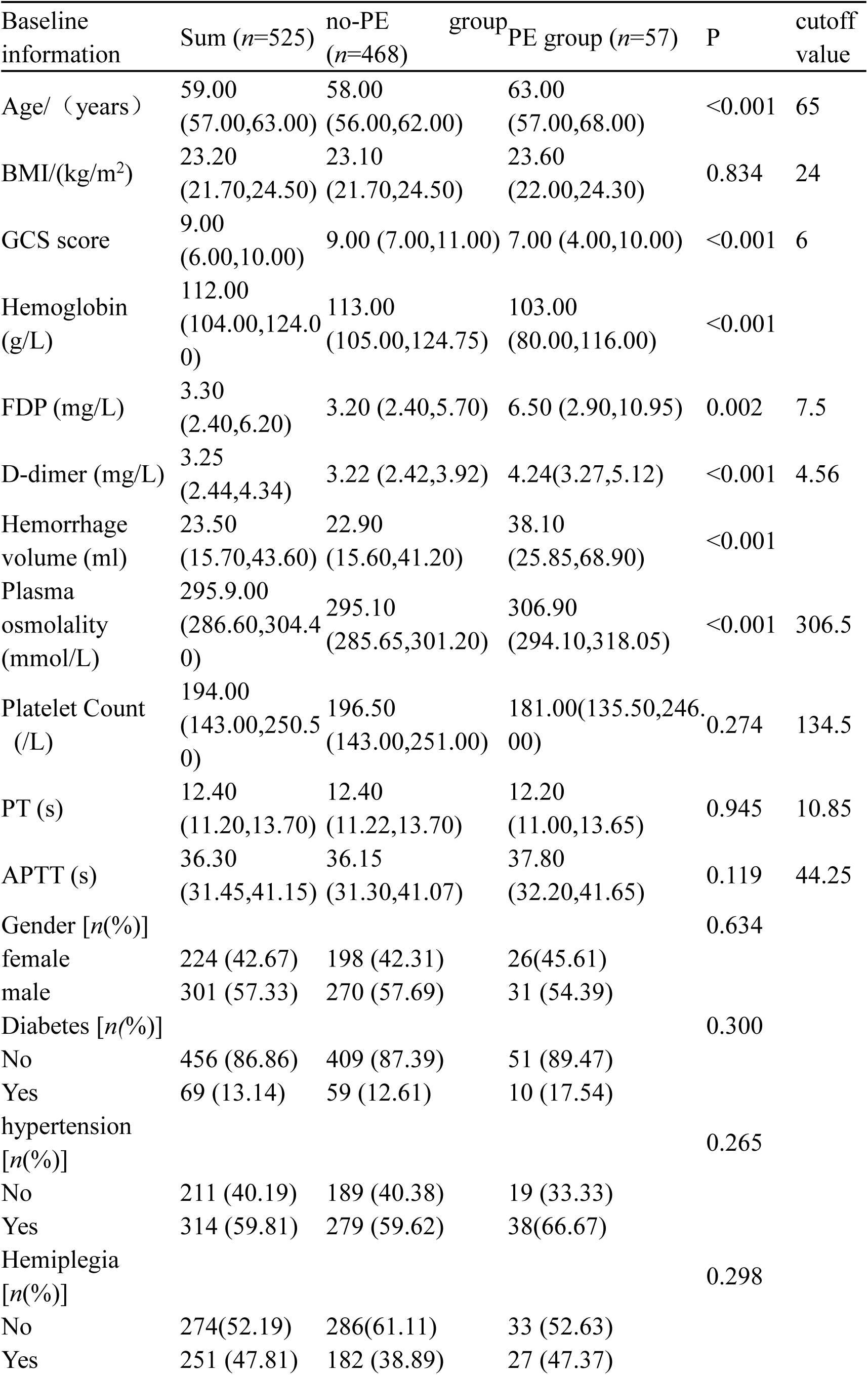
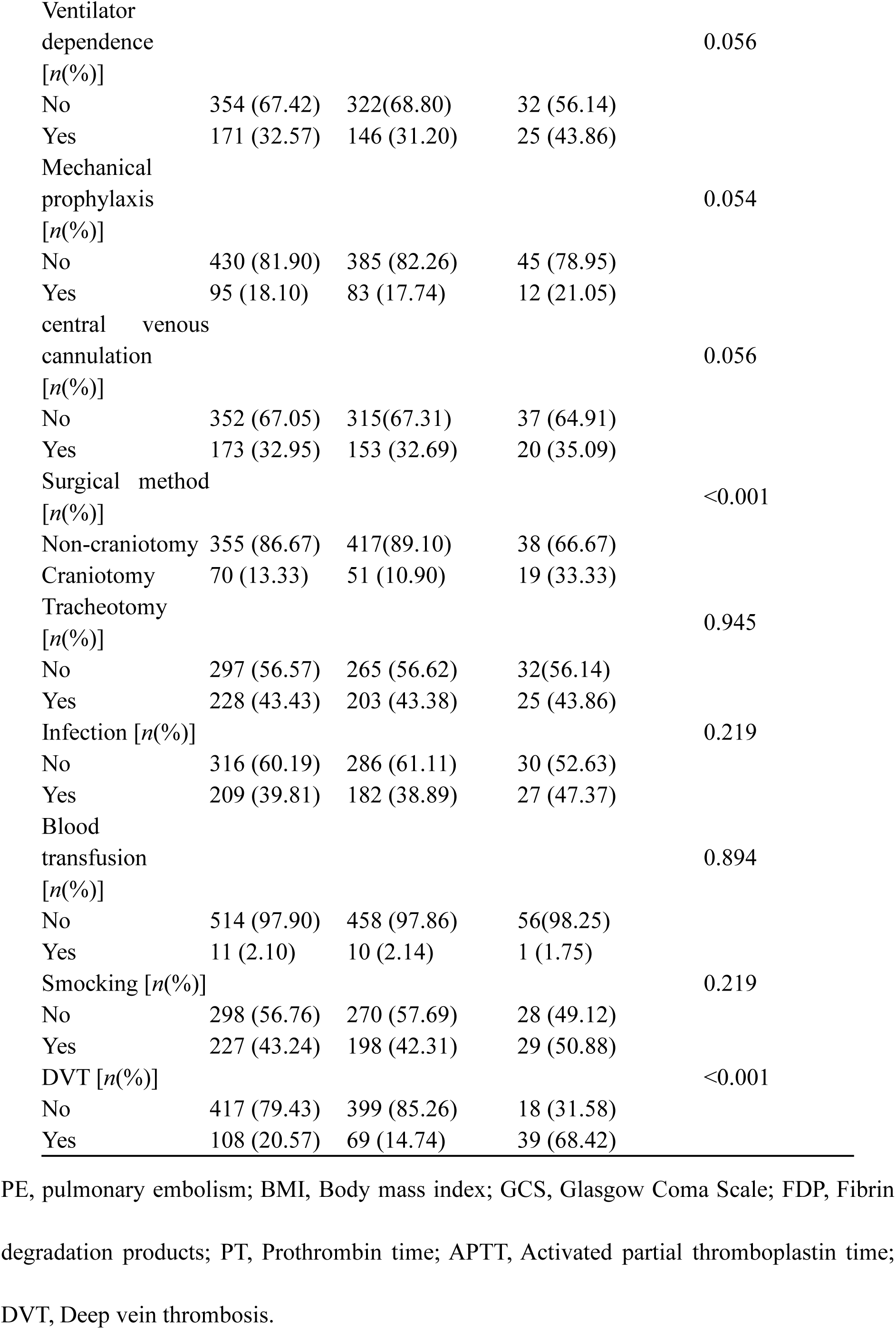
Analysis of baseline information for the training cohort [M (Q₁, Q₃)]

**Table 2.**
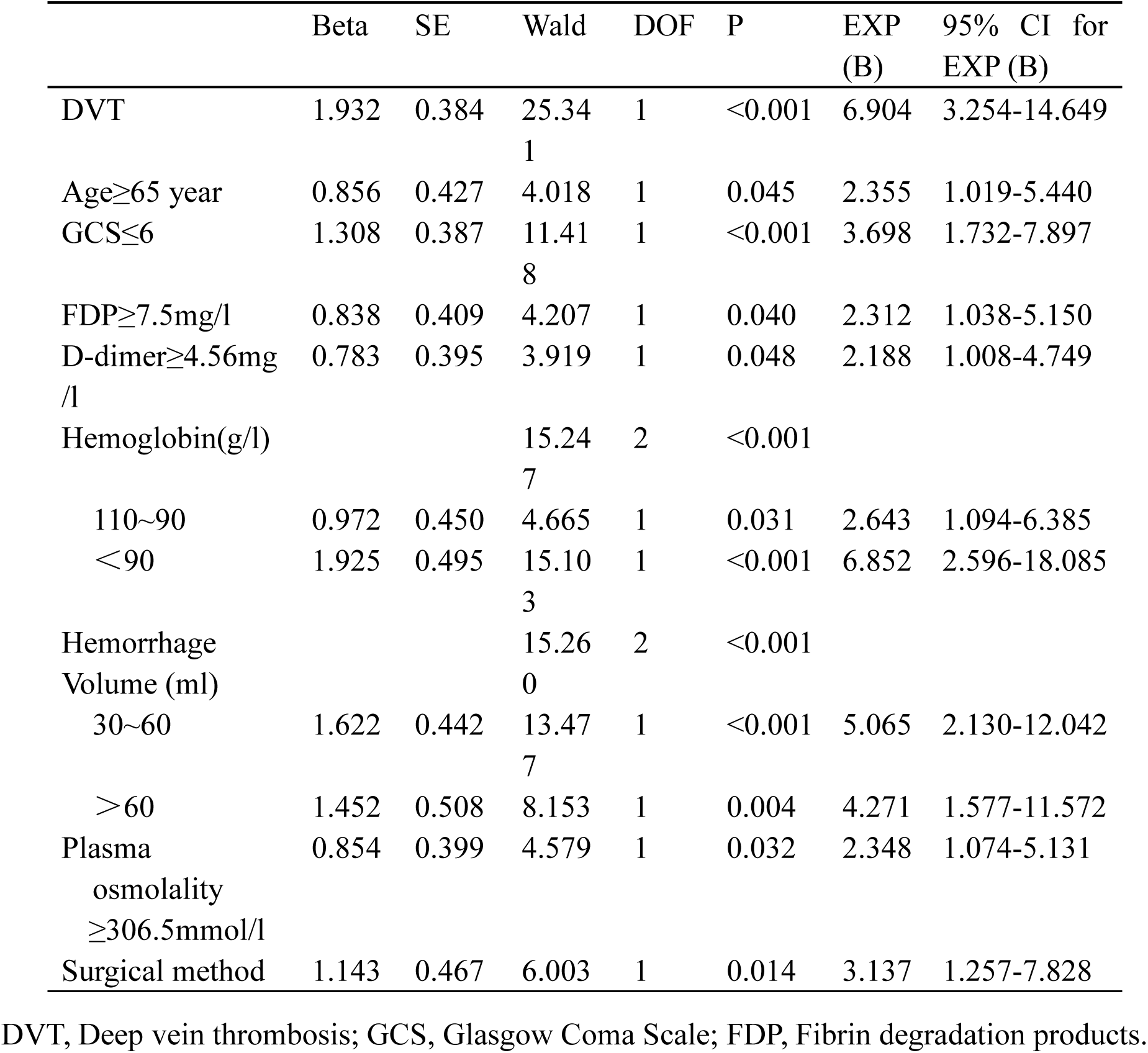
Multivariate logistic regression analysis for independent risk factors.

### Development of the Model P-P nomogram

The Figure 3A displayed the correlations between each independent risk factor and the occurrence of PE in training set. Based on the multifactorial results of the logistic regression analysis, a nomogram was established to predict the likelihood of PE after sICH surgery (Figure 3B). Each risk factor was assigned a score. The total score was calculated by adding the individual scores and locating the sum on the total-point scale axis. A vertical line was drawn downward from this point to determine the probability of PE after sICH surgery. Figure 3C illustrated the representative process of calculating the PE probability of an individual from the training set. The sICH patient had a total score of 448, he had a 59.9% probability of complicated PE.

**Figure 3.**
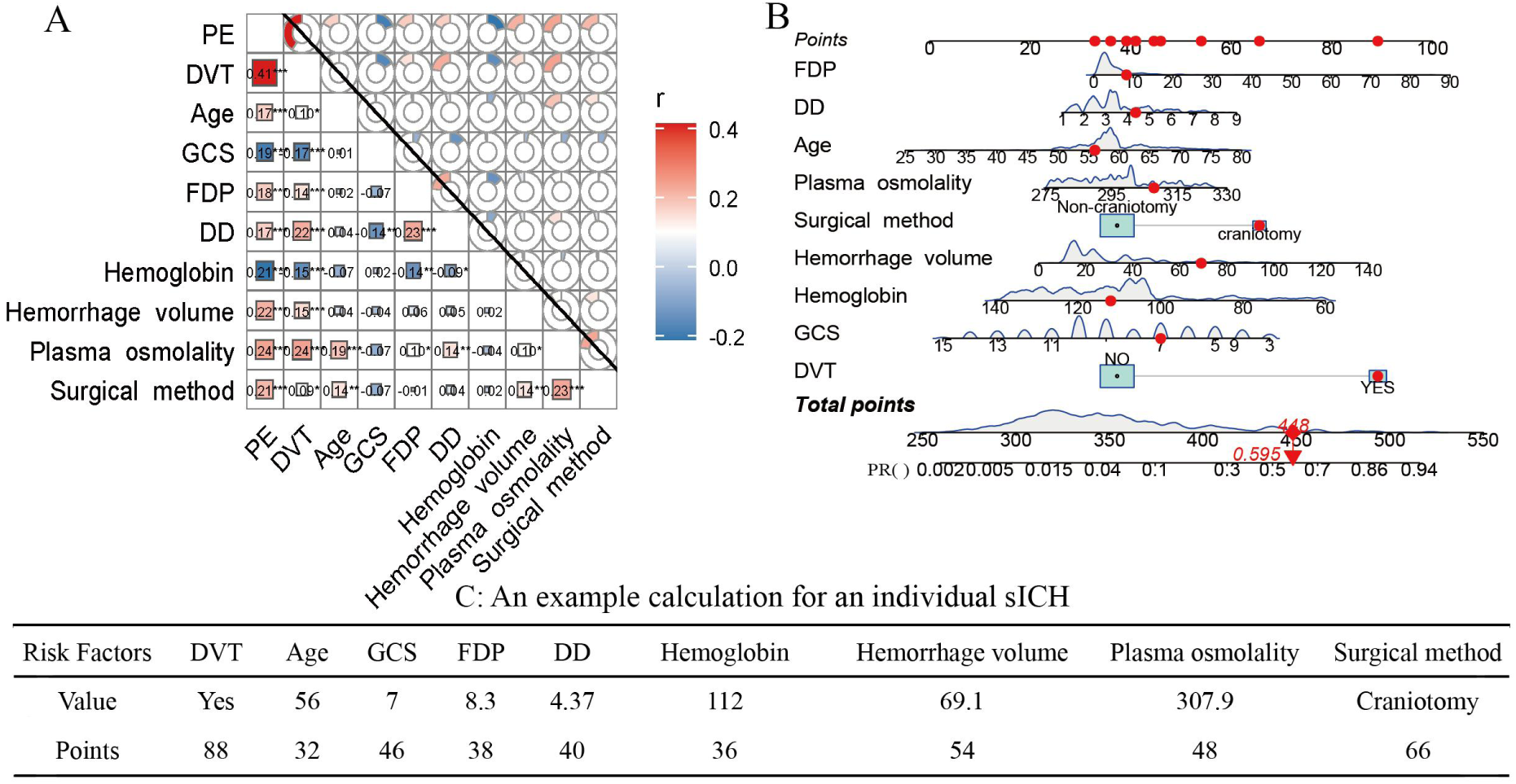
Development and illustration of Model P-P. A heatmap of the independent risk factors for PE after sICH surgery (A). DVT, Age, FDP, DD, Hemorrhage volume, Plasma osmolality and Surgical method were positively correlated with PE, while GCS and Hemoglobin were negatively correlated with PE. The nomogram (Model P-P) for predicting PE after sICH surgery (B). The representative process of calculating the PE probability of an individual from the training set (C). PE, pulmonary embolism; sICH, spontaneous intracerebral hemorrhage; DVT, Deep vein thrombosis; GCS, Glasgow Coma Scale; FDP, Fibrin degradation products; DD, D-dimer.

The AUC of Model P-P was 0.910 (95% CI: 0.857-0.962), and the model demonstrated a sensitivity of 84.2% and a specificity of 94%. The Figure 4 showed, the calibration curves of the Model P-P closely resembled the ideal curves. The clinical value of Model P-P was evaluated using DCA. The results indicated that, within the threshold risk range of 2% to 96%, the model demonstrated effectiveness.

**Figure 4.**
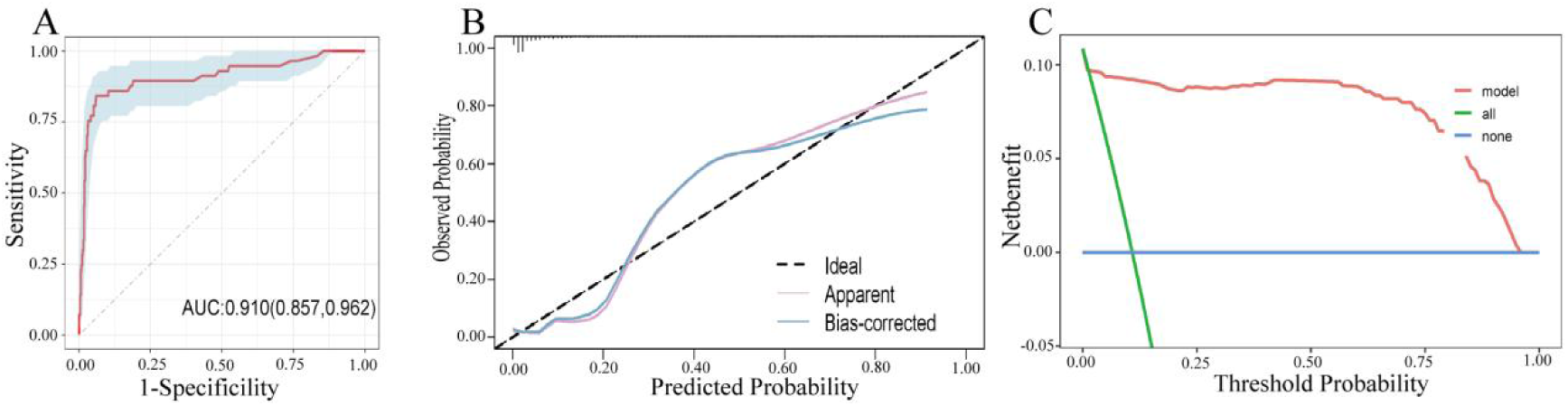
Evaluation of the Model P-P. The ROC curve (A), the calibration plot (B), and the DCA (C) of the nomogram in the training set. The above figures showed that the model’s diagnostic efficacy was deemed satisfactory and potentially offer some clinical utility. ROC, Receiver Operating Characteristic; AUC, Area Under the Curve; DCA, decision curve analysis.

### Validation of the nomogram of Model P-P

As shown by the ROC curve of the Model P-P in the internal validation set, the AUC value was 0.843 (95% CI: 0.773-0.913), and the sensitivity and specificity was 55.6% and 99% respectively. The calibration curve of the Model P-P exhibited a high level of consistence between prediction and actual observation in the validation set. DCA demonstrated that the model provided net benefits across a wide range of threshold probabilities.

The nomogram was externally validated with 247 patients, showing similar performance to that of the training set. In the external validation set, the Model P-P demonstrated an AUC of 0.945(95% CI: 0.912-0.978), a sensitivity of 92.3% and a specificity of 90.9%. The calibration curve of Model P-P in the external validation set illustrated an effective calibration performance. Figure 5F indicated the DCA revealed clinical utility within a range of 1% to 93% threshold risk.

**Figure 5.**
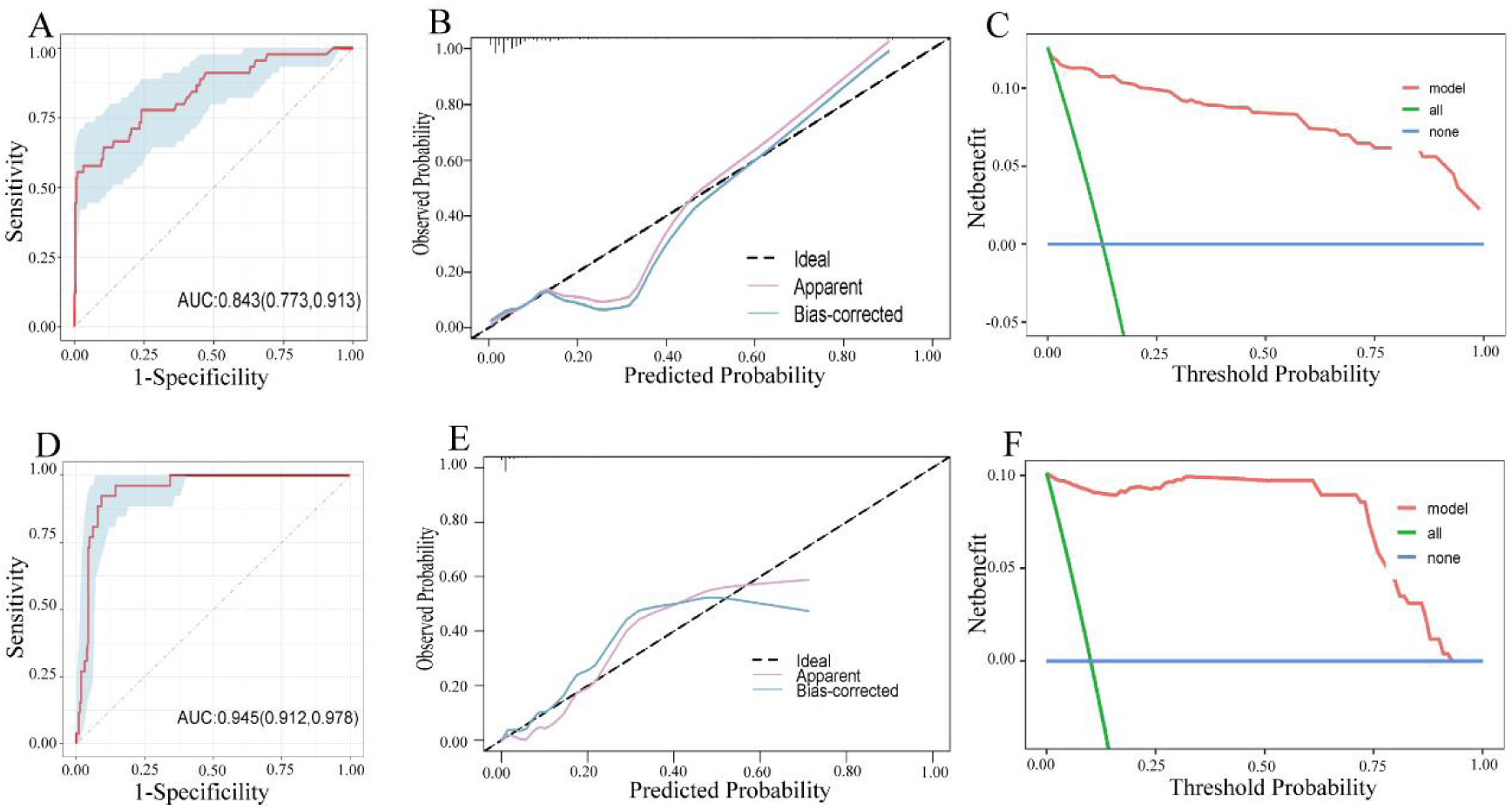
Validation of the nomogram. The ROC curve (A), the calibration plot (B), and DCA (C) of the nomogram in the internal validation set. The ROC curve (D), the calibration plot (E), and the DCA (F) of the nomogram in the external validation set. Both internal and external validations indicated that the model possessed a high level of diagnostic precision and could help healthcare professionals make reasonable decisions. ROC, Receiver Operating Characteristic; AUC, Area Under the Curve; DCA, decision curve analysis.

### Comparison with the similar model

A previous research preliminarily established a risk prediction model (Model A) for PE after sICH surgery. The additional 50 sICH patients recently admitted to our hospital were assessed separately in the Model A and Model P-P. The ROC curve was then plotted based on the total score, and the result showed the AUC of model A was 0.785 (95% CI: 0.6142-0.9552), with a sensitivity of 64.3% and a specificity of 86.1%, while the AUC of model P-P was 0.894 (95% CI: 0.8023-0.9854), with a sensitivity of 71.4% and a specificity of 88.9%. The AUC of Model A was found to be slightly lower than that of Model P-P. Figure 6B displayed the discriminatory performance of Model P-P and Model A in distinguishing complicated PE after sICH surgery.

**Figure 6.**
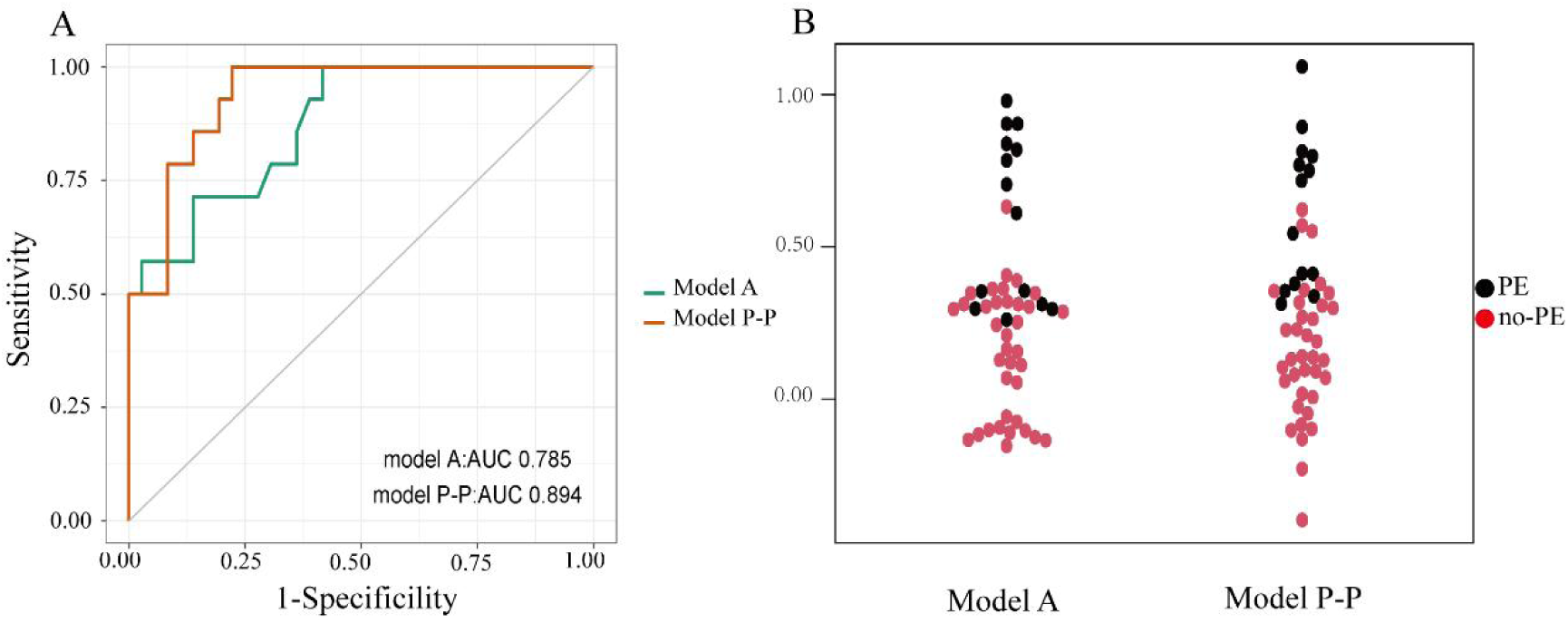
Comparison of Model P-P and Model. **A.** The AUC of Model P-P was found to be significantly higher than that of Model A (A). (P<0.05) The Bee diagrams of Model P-P and Model A (B), Model P-P had better discrimination than Mode A. PE, pulmonary embolism, ROC, Receiver Operating Characteristic; AUC, Area Under the Curve; DCA, decision curve analysis.

## Discussion

Early detection and intervention for PE were highlighted as pivotal factors influencing the outcomes of sICH.^19,20^ However, the prediction of PE after sICH surgery posed clinical challenge, attributed to the intricate pathogenesis and diverse manifestations of the condition, thereby underscoring an urgent need for more efficacious predictive instruments. Existing diagnostic modalities were not exempt from limitations, such as the potential for false positives and the necessity for specialized radiologist expertise, which could lead to diagnostic delays and inaccuracies.^21^ In view of the elevated morbidity and mortality rates linked to sICH treatments,^22^ the methods for forecasting PE after sICH surgery need further development and refinement to augment their effectiveness.

In this study, we developed a novel Model P-P for early prediction of PE after sICH surgery. It was specifically designed for critical sICH patients in neurosurgical intensive care unit (NICU). The mean duration of NICU hospitalization for sICH patients was 19 days,^23^ and complications such as DVT, PE, and infections were frequently observed.^24^ Thus, NICU-related factors could not be ignored. The range of risk factors considered in this study was broader than in previous similar research.^18,25^ We not only included the conventional risk factors for venous thromboembolism and PE, such as age, gender, BMI, etc., and the specific risk factors for sICH, such as volume of cerebral hemorrhage, GCS, surgical method, etc, but also the risk factors in the NICU, including ventilator dependence, tracheotomy, central venous cannulation, etc.

Previous research on predicting PE after sICH surgery primarily analyzed potential risk factors but without sufficient quantification.^25^ This deficiency resulted in an absence of intuitive quantitative metrics for PE prediction, and limited the applicability of prediction tool. The fundamental feature of R language is its high level of visualization, which makes the presentation of results more understandable.^26^ In this study, the advantages of the Model P-P based on R language over traditional prediction models were as follows: first, the simple and straightforward nomogram prediction model could be applied to clinical decision-making more easily than complex statistical models. Second, the nomogram prediction model would be able to predict clinical outcomes based on individual characteristics, thus promoting personalized treatment.^27^

The advantages of model P-P are not solely due to its previously highlighted strengths but also our inclusion of some specific indicators, including the surgery method and plasma osmolality. Such indicators were insufficiently stressed in the existing literatures,^18 25^ urging for a more comprehensive consideration in the development of predictive tools for PE. DVT is one of the most common postoperative complications, especially after craniocerebral surgery.^28^ First, brain tissue and cerebral vascular damage during surgery may lead to the release of various tissue factors and cause brain retraction injuries.^29^ Second, craniotomy, associated with increased blood loss and hemodynamic instability, could slow venous blood flow.^30^ Third, the extended duration of anesthesia and surgery limited patient mobility, resulting in diminished circulation in the lower extremities contribute to blood stasis.^31,32^ In general, these surgery-related factors mentioned above, facilitated thrombus formation and the incidence of PE. In the univariate and multivariate logistic regression analyses conducted, surgical method was observed statistical significance. The Model P-P revealed a predicted score of 66 points for craniotomy and 38 points for non-craniotomy. The results of our study indeed demonstrated that craniotomy was associated with an increased risk of PE after sICH surgery.

The hypercoagulable state of blood was recognized as a critical factor in PE formation^33^ and plasma osmotic pressure was confirmed as a reliable indicator of hypercoagulability.^34^ Consequently, plasma osmotic pressure was utilized in this study to examine its role in the hypercoagulable state associated with the development of PE. This study revealed that a plasma osmolality exceeding 306.5 mmol/L was identified as a significant risk factor for the occurrence of PE after sICH surgery. This discovery presents a novel potential biomarker for clinical practice.

Profiting from all the contributive factors mentioned above, as shown in the results, the Model P-P exhibited satisfying performance in both internal and external validations. The calibration plot demonstrated a high level of consistence between prediction and actual observation, rendering Model P-P clinically beneficial. Furthermore, when compared to similar prediction model for PE after sICH surgery, the Model P-P showed superior accuracy and practicality, with statistically significant differences. Consequently, this model seems to possess higher practical value and better comprehensive performance in predicting PE after sICH surgery.

Although this study yielded promising preliminary results, it was important to admit its limitations. First, this study was limited by its single-center design and small sample size, which could introduce statistical biases and restrict its generalizability. Second, although the prediction model showed plausible efficacy in both internal and external validations within this study, further external validation across multiple centers is still required. Third, the expansion using of Model P-P in other diseases is restricted without some certain design modifications and parameter adjusting.

## Conclusions

This study had developed a quantitative, visualized and convenient nomogram tool for early prediction and assessment of PE in sICH patients. It demonstrated a validated favorable predictive accuracy in clinical applications, and may assist in making personalized treatment for sICH patients in NICU.

## Data Availability

The datasets generated and analyzed during the current study are not publicly available due privacy /ethical restrictions but are available from the corresponding author on reasonable request.

## Acknowledgments

Thanks to the First Affiliated Hospital of Chongqing Medical University for providing data support.

## Abbreviations

APTT: Activated partial thromboplastin time
AUC: Area under the curve
BMI: Body mass index
CTPA: Computed tomographic pulmonary angiography
DCA: Decision curves analysis
DD: D-dimer
DVT: Deep vein thrombosis
FDP: Fibrin degradation products
GCS: Glasgow Coma Scale
NICU: neurosurgical intensive care unit
PE: Pulmonary embolism
PT: prothrombin time
ROC: Receiver operating characteristic
sICH: Spontaneous intracerebral hemorrhage

